# Changes in the adult consequences of adolescent mental health: findings from the 1958 and 1970 British birth cohorts

**DOI:** 10.1101/2020.07.17.20156042

**Authors:** Ellen Jo Thompson, Marcus Richards, George B. Ploubidis, Peter Fonagy, Praveetha Patalay

**Affiliations:** UCL Centre for Longitudinal Studies, Department of Social Science, University College London, UK; MRC Unit for Lifelong Health and Ageing at UCL, University College London, UK; Research Department of Clinical, Educational and Health Psychology, University College London, UK

## Abstract

**Objective:** To investigate whether the associations between adolescent externalising and internalising mental health difficulties and mid-life health and social outcomes are changing across cohorts.

**Methods:** Data from 31,349 participants followed as part of two prospective national birth cohort studies: 1958 National Child Development Study (n=16091) and the 1970 British Cohort Study (n=15258). Adolescent mental health at age 16 years was assessed using the Rutter scales and operationalised as both a traditional 2-factor internalising and externalising and as a hierarchical bi-factor. Associations between adolescent psychopathology and health and wellbeing (mental health, general health, life satisfaction) and socio-economic (cohabitation, voting behaviour, education and employment) outcomes at ages 42 are estimated and illustrated across cohorts.

**Results:** The prevalence of adolescent mental health difficulties increased between those born in 1958 and 1970. Their associations with different midlife outcomes became more severe or remained similar in the 1970 compared the 1958 cohort. For instance, a stronger association with adolescent mental health difficulties was found for those born in 1970, compared with those born in 1958 for midlife psychological distress (OR 1970=1.82 [1.65;1.99], OR 1958=1.60 [1.43;1.79], cohabitation (OR 1970=0.64 [0.59;0.70], OR 1958=0.74 [1.43;1.79]), and professional occupations (OR 1970=0.75 [0.67;0.84], OR 1958=1.05 [0.88;1.24]). A hierarchical bi-factor approach to modelling adolescent mental health indicated that the associations of externalising symptoms with later outcomes were mainly explained by their shared variance with internalising symptoms.

**Conclusion:** A strengthening trend in the associations of adolescent mental health with adverse mid-life outcomes across two generations highlights the individual and societal burden of adolescent mental health problems might be increasing. If this trend continues to apply to more recently born cohorts, the wide-ranging implications of the increasing prevalence in adolescent mental health difficulties currently observed should be recognised and increased public health efforts to minimise adverse outcomes are needed.

## Introduction

Common mental health problems, such as anxiety and depression are a leading cause of disease burden.^1^ Estimated high prevalence of common mental health problems in children and adolescents is of particular concern as early onset symptoms often persist into adulthood,^2,3^ causing substantial health and social costs in adult life.^4,5^ Mental ill-health frequently occurs in childhood and adolescence, with 1 in 8 5-19 year olds with a diagnosable disorder and a higher proportion of young people reporting high levels of internalising and externalising symptoms.^6,7^ It has also been observed that the prevalence of mental ill-health symptoms is changing across generations, with twice as many young people in the UK reporting frequent feelings of depression and anxiety from 1986 to 2006,^8^ and almost twice as many adolescents experiencing high depressive symptoms in 2015 compared to 2005.^9^

Studies providing evidence for the association between adolescent mental ill-health and psychosocial adult outcomes have been recently reviewed,^10,11^ highlighting that adolescent mental ill-health has negative associations with several domains in adulthood, including socio-economic outcomes and physical and mental health. However, it is unclear whether the magnitude of these associations are also changing across cohorts. Investigating this may help inform health policy and planning in relation to the consequences and burden associated with increasing trends in adolescent mental health problems. Studies of adulthood outcomes in relation to adolescent mental health have been conducted using the British birth cohorts,^4,5,12^ but, to our knowledge no investigations of differences between cohorts has been undertaken to-date, possibly due the requirement of a sequence of long running longitudinal studies to do so. A recent study investigated the association between childhood mental health difficulties at age 7 and adolescent outcomes (14-16 years) in three cohorts their findings indicate that these associations are similar magnitude across cohorts and might be getting slightly stronger for some outcomes in recent generations.^13^

In adolescent mental health, common difficulties are usually represented in two domains, internalising and externalising symptoms, which exhibit above chance levels of co-morbidity and have common antecedents.^14^ Regarding adult outcomes, there is some evidence that externalising difficulties might have stronger negative associations with educational and economic outcomes.^5,12^ However, it is unclear whether this is still the case when the shared variance with internalising symptoms is accounted for. In recent years, to account for their comorbidity and permit investigation of overall levels of mental health difficulties, mental health has been conceptualised as a general psychopathological construct that reflects shared variance between internalising and externalising symptoms.^15,16^ In young people this general psychopathology factor explains between 70-80% of the variance in symptomology, and is highly predictive of future outcomes.^17,18^ In this study, we investigate how the construct of general psychopathology predicted later outcomes, and compare findings to a more traditional 2-factor internalising and externalising model of mental health.

The current study aimed to investigate whether cohort moderated the strength of associations between adolescent psychopathology (at age 16) and a wide-range of health and social outcomes in mid-life (at age 42) using simultaneous analysis of identically measured exposure, outcomes and controls across the 1958 National Child Development Study and the 1970 British Cohort Study. In addition, adolescent mental health was conceptualised as traditional 2-domains (internalising and externalising difficulties) and also using a more recent bi-factor general psychopathology approach. A range of health and socio-economic outcomes in midlife including mental health, general health, life satisfaction, cohabitation, voting, education attainment and employment type were investigated to provide a comprehensive picture of the health and socioeconomic outcomes related to adolescent mental health and whether these are changing across cohorts.

## Methods

### Participants

This study used data from 31349 participants from two British birth cohort studies.

### 1958 NCDS

The 1958 National Child Development Study (1958 NCDS) follows the lives of 17,415 people born in England, Scotland and Wales in a single week of 1958.^19^ Since the birth survey in 1958, there have been 10 further surveys of all cohort members at ages 7, 11, 16, 23, 33, 42, 44, 46, 50 and 55 years. In 2000 at age 42 years, 11,419 study members (50.7% female) participated in the survey. The analytic sample for this study consisted of 16091 participants (92.39% of the original sample) having excluded those who had died or emigrated by age 42 years.

### 1970 BCS

The 1970 British Cohort Study (1970 BCS) follows the lives of 17198 people born in England, Scotland and Wales in a single week of 1970.^20^ Since the birth survey in 1970, there have been eight surveys at ages 5, 10, 16, 26, 30, 34, 38 and 42 years. In 2012 at age 42 years 9841 study members (52.20% female) participated in the survey. The analytic sample for this study consisted of 15258 participants (88.72% of the original sample) having excluded those who had died or emigrated by age 42 years.

### Measures

#### Mental health at age 16

At age 16 one parent (usually the mother) of both 1958 NCDS and 1970 BCS cohort members completed the Rutter A scale,^21^ consisting of 18 symptoms for which the parent was asked to indicate whether each ‘does not apply’ ‘applies somewhat’ or ‘definitely applies’. Confirmatory 2-factor and bi-factor analyses were applied to derive latent factor scores, which were treated as continuous measures of internalising and externalising symptoms and general psychopathology. We tested the measure for invariance between cohorts and found it to be invariant at the configural, metric and scalar levels, indicating between cohort comparisons could be made using this measure (see eTable 1).^22^

**Table 1.** Means / proportions and 95% confidence intervals (CIs) for exposure and outcome variables.

|  |  |  | **Cohort** |  |
| --- | --- | --- | --- | --- |
|  |  |  | **1958 NCDS** | **1970 BCS** |
|  |  |  | *n* = 16,091 | *n* = 15,258 |
|  |  |  | Mean / % (95% CIs) | Mean / % (95% CIs) |
| **Exposure**<br>**(age 16)** | Mental Health (2-factor model) | Externalising Symptoms | Mean = .06 (.056; .074) | Mean = .24 (.23; .25) |
|  |  | Internalising Symptoms | Mean = .05 (.04; .06) | Mean = .12 (.12; .13) |
|  | Mental Health (Bi-factor model) | Externalising Symptoms | Mean = .07 (.06; .07) | Mean = -.18 (-.18; -.17) |
|  |  | Internalising Symptoms | Mean = .02 (.02; .03) | Mean = .07 (.06; .08) |
|  |  | General psychopathology | Mean = .07 (.07; .08) | Mean = .22 (.21; .23) |
| **Outcome**<br>**(age 42)** | Psychological distress (continuous score) | Malaise inventory | Mean = 1.54 (1.51; 1.58) | Mean = 1.92 (1.87; 1.96) |
|  | High psychological distress (binary) | Malaise inventory >3 | % = 11.9 (11.3; 12.4) | % = 16.7 (16.1; .17.04) |
|  | General Health |  | Mean = .02 (-.01; .03) | Mean = .03 (.01; .05) |
|  | Life Satisfaction |  | Mean = 7.25 (7.22; 7.29) | Mean = 7.31 (7.27; 7.34) |
|  | Living with spouse or partner | No | % = 20.1 (19.3; 21.6) | % = 25.5 (24.5; 26.4) |
|  | Voted in the last general election | No | % = 23.8 (22.9; 24.6) | % = 27.7 (26.4; 29.1) |
|  | Obtained a degree | No | % = 80.8 (80.2; 81.5) | % = 69.0 (68.1; .69.9) |
|  | Employment type | Professional/managerial | % = 57.6 (56.7; 58.6) | % = 58.7 (57.7; 59.8) |

#### Health and psychological outcomes at age 42

At age 42 psychological distress was measured in both cohorts using the 9 item version of the Malaise Inventory,^23^ which includes items assessing presence of symptoms such as ‘feel miserable and depressed’, ‘get worried about things’ and ‘get easily upset or irritated’. On the continuous sum score, a high score indicates greater psychological distress and there is an established cut-off of a score of above 3 indicating high psychological distress.

Life satisfaction was assessed by the question ‘How had life turned out so far’: on a scale of 0 to 10, where 0 is not at all satisfied and 10 is completely satisfied. General state of health was also measured in both cohorts at age 42. In the 1958 NCDS cohort members were asked to rate their general health using 4 categories (1= Excellent; 2 = Good; 3 = Fair; 4 = Poor); and in the 1970 BCS cohort members were asked to rate their general health using 5 categories (1= Excellent; 2 = Very good; 3 = Good; 4 = Fair; 5 = Poor). To harmonise this variable, each measure was normalised.

#### Social and economic outcomes at age 42

Whether the cohort member lives with a spouse or partner, their highest academic qualification, their employment status (managerial / professional / non-managerial / professional / never worked / long-term unemployed), and whether the cohort member voted in the last general election was recorded in both cohorts at age 42. A binary variable was derived for highest academic qualification, which represented those who had a degree (coded 1) vs those who had no or other qualifications (coded 0). A binary variable was also derived for employment status which represented those in professional managerial positions (coded 1) vs those in any other employment status.

#### Covariates

Socio-economic and health covariates derived from birth and early childhood were included in the analysis. All covariates were measures identically between cohorts. Covariates derived from birth and early childhood included birth weight, maternal smoking during pregnancy, whether cohort members were breastfed, childhood cognition and childhood body mass index. Covariates derived from parents included paternal social class at age 10/11 years (higher managerial, administrative and professional occupations, intermediate occupations, routine and manual occupations), whether the mother of the participants was employed (birth to age 5 years), divorced (by age 10 or 16 years), and years of education of mother and father.

### Statistical Analysis

#### Factor Models

Measurement models were run on the age 16 mental health measure. Confirmatory Factor Analyses (CFA) were conducted to test two models. A 2-factor model was estimated which confirmed the factor structure for internalising and externalising psychopathology. Second, a bi-factor model where each symptom loads on to general psychopathology as well as internalising and externalising factors. CFA models were estimated using Mplus 8^24^ and the weighted least square means and variances (WLSMV) estimator was used for categorical manifest variables.^25^ Measurement invariance of the age 16 Rutter measure between cohorts was tested to establish comparability of the measure between the cohorts (and measures were found to be invariant, details in supplementary file).

To compare the relative variance explained of both the general and specific factors, the explained common variance (ECV) for each factor was calculated for externalising, internalising and general psychopathology. ECV values range from 0 to 1, with values closer to 1 suggesting a greater share of variance explained. There are no established cut-offs in ECV values; it has been suggested that ECV values ranging from .60 to .85 indicate that a factor is the main source of shared variance.^26^

#### Missing data

Multiple imputation using chained equations was implemented to impute missing data with fifty imputations conducted separately in both cohorts (see eTable 4 for counts and percentages of non-imputed data for each predictor and outcome). This multiple imputation approach includes a number of covariates and auxiliary variables in the models, which maximises the plausibility of the missing at random (MAR) assumption.^27^

#### Regression analyses

Linear and logistic multivariate regression models were fitted to test associations between psychopathology at age 16 and health and socio-economic outcomes at ages 42, adjusting for a rich set of early life covariates. Model comparison tests between models with and without cohort interactions were conducted and given significant improvements in model fit when including cohort*age 16 mental health interaction terms, stratified models are presented by cohort. We illustrate the cohort differences in *exposure-outcome associations* across the distribution of the exposure using predictive margins from the regression models. We also investigated whether associations between adolescent mental health and midlife outcomes differed between sexes using age sex*cohort*age 16 mental health interactions. An alpha level of .05 was used with adjustment made for rate of false discovery due to multiple outcomes testing using the Benjamini-Hochberg procedure.^28^

## Results

### Factor models

The first CFA estimated the 2-factor model using separate externalising and internalising latent factors. Factor loadings and model fit statistics for this model are displayed in supplementary table S1. The second CFA estimated the bi-factor model with externalising, internalising specific and a general psychopathology bi-factor. Factor loadings and model fit for this model are displayed in supplementary table S2. Estimating the ECV indicates that the general psychopathology factor accounted for 72.1% of the variance in symptoms, externalizing symptoms accounted for 14.8% of the variance and internalising symptoms accounted for 13% of the variance.

### Descriptive statistics

Table 1 displays the mean factor scores for adolescent mental health and the means / proportions for each harmonised outcome variable. Most mental health symptoms scores at ages 16 were higher for in the 1970 cohort than for those 1958 cohort (with the exception of the externalising specific factor from the bi-factor model). eTable 5 displays the correlations between adolescent mental health and age 42 health and social outcomes. Correlations are similar or of higher magnitude between each exposure-outcome pair for those born in the 1970 cohort than for those in the 1958 cohort.

### Associations between adolescent mental ill-health and health and social outcomes in adulthood

Model comparison tests of models with and without cohort interactions indicated that for all outcomes models with cohort interactions with age 16 mental health from the bi-factor model (and all except two from the 2-factor model) had improved model fit than without the cohort interactions, supporting the presentation of cohort stratified estimates (log-likelihood tests for the model comparisons are presented in Table 2).

**Table 2.** Likelihood ratio tests comparing models with main effects only and cohort interactions to assess cohort differences in associations between age 16 mental health exposures and age 42 outcomes

|  |  | Log likelihood difference test |  |  |  |  |  |  |  |
| --- | --- | --- | --- | --- | --- | --- | --- | --- | --- |
|  |  | Psychological distress | High psychological distress (Yes) | General Health | Life Satisfaction | Living with spouse or partner (Yes) | Voted in the last general election (Yes) | Obtained a degree | Employment type (professional/managerial) |
| Models compared | | $\chi^2$ (df) | $\chi^2$ (df) | $\chi^2$ (df) | $\chi^2$ (df) | $\chi^2$ (df) | $\chi^2$ (df) | $\chi^2$ (df) | $\chi^2$ (df) |
| Main effects VS Cohort*Age 16 mental health | Two factor model | **27.14 (2)** | **7.98 (2)** | **18.18 (2)** | **19.72 (2)** | **27.97 (2)** | **10.09 (2)** | 2.49 (2) | 1.57 (2) |
|  | Bi-factor model | **185.58 (4)** | **102.40 (4)** | **14.80 (4)** | **40.76 (4)** | **89.25 (4)** | **51.43 (4)** | **554.63 (4)** | **21.04 (4)** |
| Cohort*Age 16 mental health VS Sex*cohort*Age 16 mental health | Two factor model | 2.58 (4) | **12.21 (4)** | 4.82 (4) | 0.81 (4) | **15.41(4)** | 2.34 (4) | 4.74 (4) | **10.45 (4)** |
|  | Bi-factor model | 4.77 (6) | **14.28 (6)** | 10.23(6) | 1.47 (6) | 12.12 (6) | 8.88 (6) | 5.15 (6) | 9.67 (6) |

Note: Results in bold are where log-likelihood difference test indicated differences in models being compared (p<0.05)

All coefficients for the exposure-outcome associations stratified by cohort are displayed in Table 3.

**Table 3.**
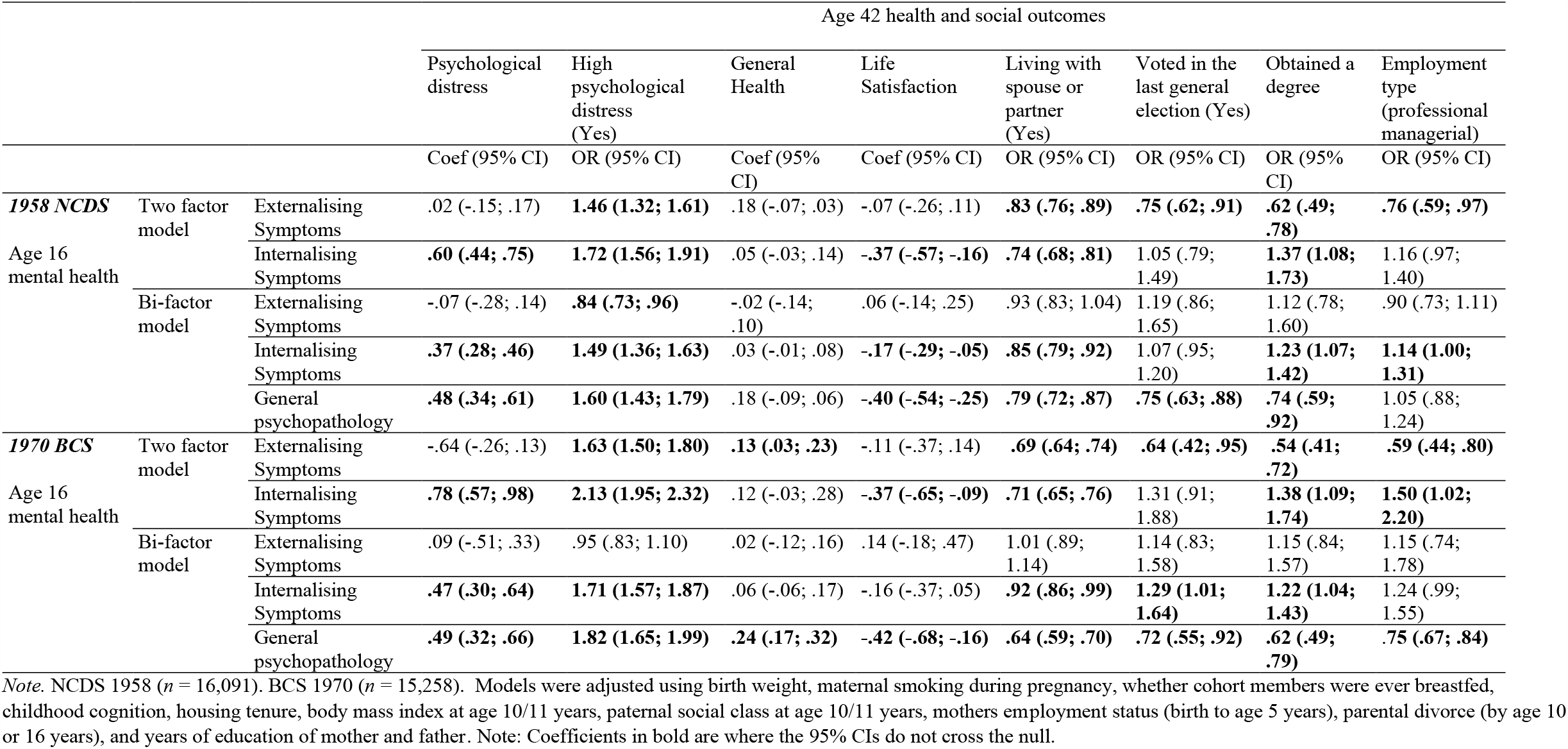
Multivariate regressions estimating associations between adolescent psychopathology and age 42 health and social outcomes in NCDS 1958 and BCS 1970 cohorts

Externalising symptoms (from the 2-factor model) were associated with high psychological distress and lower likelihood or living with a partner, low education attainment and lower occupational class in both cohorts, with consistently poorer associations in BCS 1970. Internalising symptoms (from the 2-factor model) were associated similarly in both cohorts with life satisfaction, living with a spouse and obtaining a degree in both cohorts. However, a clear cohort effect is observed for age 42 psychological distress where higher internalising symptoms at age 16 is associated with 1.72 higher odds (95% CI: 1.56;1.91) in NCDS1958 and 2.13 higher odds (95%CI: 1.95;2.32) (eFigure 1 illustrates associations by cohort for each 2-factor exposure-outcome pair).

**Figure 1.**
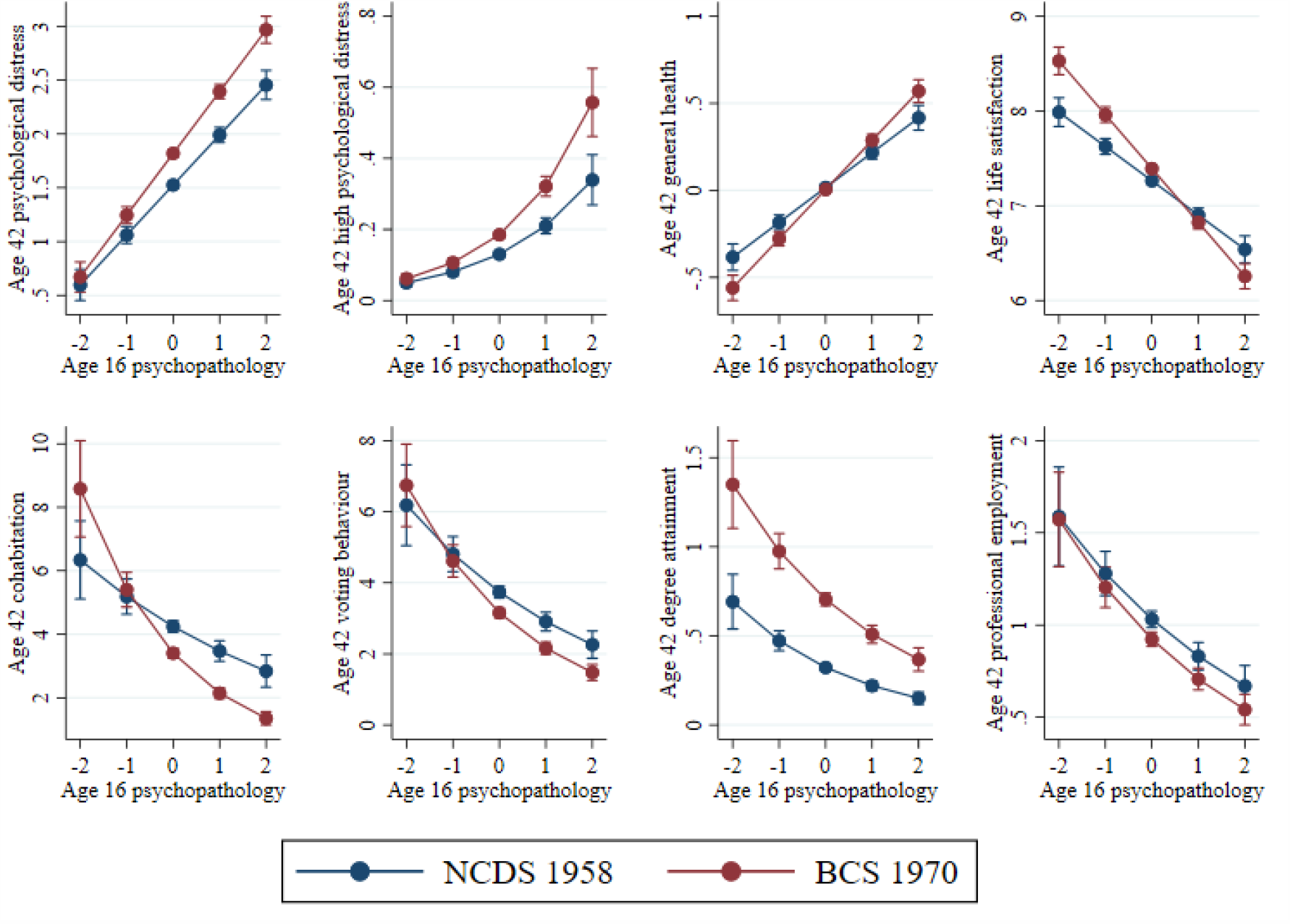
Predicted probabilities for mental health at age 16 (x-axis) association with each outcome at age 42 in the two cohorts.

General psychopathology at age 16 was associated with poorer outcomes for most adult outcomes examined in both cohorts. Figure 1 illustrates the associations by cohort for general psychopathology and each outcome. We observe that at higher levels of adolescent difficulties, for several outcomes, including psychological distress, general health, higher education attainment, and employment, the associations with adverse adult outcomes are greater in the BCS 1970 cohort. For instance, age 16 mental health is not associated with lower likelihood of higher occupational class in the NCDS 1958: OR=1.05 (.88;1.24) whereas it is associated with substantially lower odds of higher occupational class in the BCS 1970 cohort: OR=0.75 (0.67;0.84).

After accounting for the shared variance associated with general psychopathology, there were no associations between externalising symptoms (from the bi-factor model) and any of the outcomes tested in both cohorts. Internalising symptoms (from the bi-factor model) were still independently associated with several adulthood outcomes including greater psychological distress, lower life satisfaction, and lower likelihood of cohabitation; however, the specific internalising symptoms were associated with attaining a higher degree. Supplementary figure eFigure 2 illustrates the associations by cohort for externalising and internalising bi-factor exposure-outcome pair.

**Figure 2.**
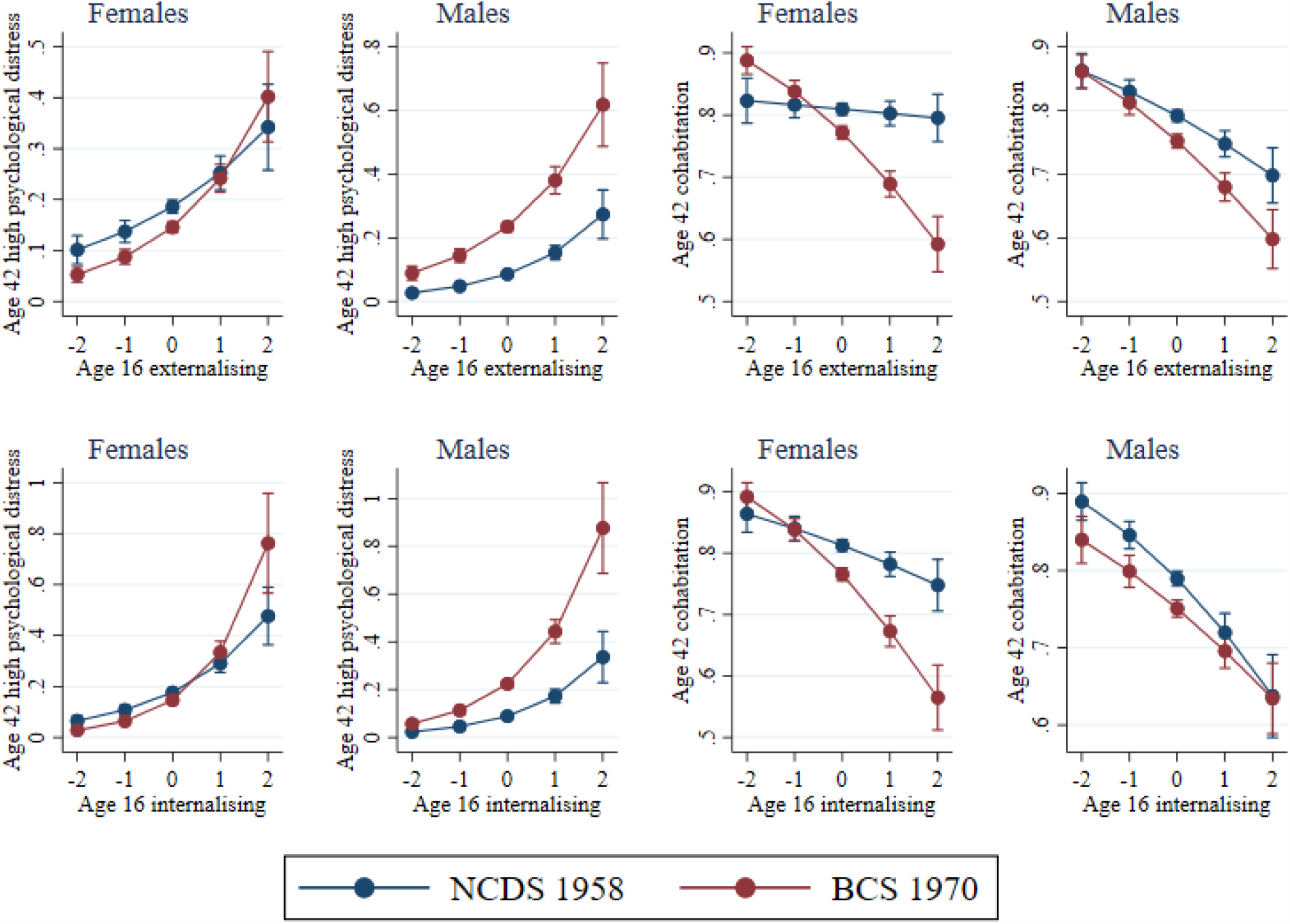
Predicted probabilities in males and females for mental health at age 16 (x-axis) association with high psychological distress and cohabitation at age 42 in the two cohorts

We also conducted log-likelihood tests with models including sex*cohort interactions to examine whether the exposure-outcome associations across cohorts were moderated by sex (Table 2). For most exposure-outcome associations, the models including sex interactions did not have better model fit, indicating that cohort differences observed are similar for males and females. A consistent exception was psychological distress at age 42 years, where we observe that the cohort effect is starker in males with either externalising and internalising mental health at age 16 (from 2-factor model) and the bi-factor (see Figure 2 and eFigure 3). For likelihood of cohabitation we see that the lower likelihood by greater age 16 mental health difficulties are similar in males and females in the 1970 cohort, however, in the 1958 cohort this association was not observed in females and lower likelihood of cohabiting was only observed in males (see Figure 2) and there is evidence of cohort effects being moderated by sex for professional occupations (see eFigure 4).

## Discussion

The findings of the paper highlight that the consequences of early life mental ill-health on a wide range of mid-life outcomes became more severe or remained similar in a cohort born in 1970 compared to a generation prior born in 1958. Alongside growing prevalence of adolescent mental ill-health, which we also observe between these two cohorts, the larger mental health based inequalities in life outcomes in the more recently born cohort call attention to the large and increasing public health and societal burden of mental ill-health through the life course.

The associations between early life mental health and mid-life social and economic outcomes followed a consistent general pattern of direction of effects, however, the magnitude of these associations became stronger in the younger generation for certain outcomes predicting greater mid-life mental health, lower likelihood of co-habitation with a partner, likelihood to vote and being in professional employment. For instance, greater general psychopathology at 16 was associated with a 60% increased risk of mid-life psychological distress in the 1958 cohort and 82% increased risk in the 1970 cohort. In another example, adolescents with higher symptoms were 21% less likely to be in cohabiting relationship at age 42 in the 1958 born cohort, whereas they were 36% less likely to be co-habiting in the 1970 born cohort. If sustained in the long term in more recently born cohorts (e.g. Millennium Cohort Study, born 2000-01), a hypothesis we cannot currently test with the available data, this trend will likely have a considerable societal and public health impact.

Some cohort effects were also gendered, and we observe starker cohort differences in males in relation to their mid-life psychological distress and starker cohort differences in females in relation to their likelihood of being in a partnership in mid-life. Reasons for this gendered pattern need better understanding and in regard to mental health outcomes, might reflect greater stigma faced by males preventing them for help seeking despite a greater number experiencing mental health difficulties.^9^

With respect to how adolescent mental health was conceptualised, internalising symptoms were associated with higher psychological distress, a higher likelihood obtaining a degree, and poorer life satisfaction at age 42 years. This also remains the case for most outcomes (excluding life satisfaction), when using a specific internalising factor accounting for covariance with externalising symptoms in the bi-factor model. Externalising difficulties were associated with poorer general health, voting behaviour and degree attainment; however, this was no longer the case when accounting for the covariance between internalising and externalising symptoms using the bi-factor model. The findings from the 2-factor model of internalising and externalising symptoms are in line with previous literature which shows both domains are associated with poorer outcomes.^5,10–12^ Notably however, the bi-factor approach which allows us to account for the high levels of shared variance between these domains, highlights that most of the negative associations are predicted by the general psychopathology factor. This finding contradicts previous evidence from a longer term longitudinal birth cohort study, which indicated externalising symptoms as an important factor in the prediction of mid-life socio-economic outcomes, while controlling for internalising difficulties.^5^ However, the former study was based on data from the 1946 British birth cohort, where symptoms were reported by teachers and externalising and internalising have weak associations, whereas the findings from this study are based on parent report of symptoms, where internalising and externalising symptoms were highly correlated.

### Strengths and limitations

Successive national population-representative birth cohort studies with comparable measures of common mental health problems provide a unique opportunity to make robust comparisons across cohorts using aggregated analytical techniques. Previous cross-cohort research is limited by differences in how outcomes and exposures are assessed,^13^ resulting in uncertainty of whether observed cohort effects are method artefacts. By using completely harmonised exposure, outcome and control variables we can be confident that the cohort effects we observe are robust, especially since the mental health measures showed evidence of measurement invariance across cohorts. Another notable strength of this study was the range of outcomes across health and socio-economic domains. One limitation of the data was that rates of attrition across cohorts were not identical. To address the issue of missing data that is unavoidable in long running birth cohorts, multiple imputation was applied with a detailed set of early life auxiliary predictors to minimise the risk of observed effects being due to potential differences in those retained in the cohorts.

### Implications

Recent research highlights growing levels of adolescent internalising symptoms in more recently born cohorts.^9^ This is also observed in the cohorts compared in our study, with those born in 1970 reporting greater levels of mental health difficulties in adolescence and midlife.^29^ One often suggested explanation for observed increases in adolescent mental health problems across cohorts is that it might be driven by differences in how these measures are completed due to less stigma and more awareness around mental health.^30^ However, we establish that there is measurement invariance for the Rutter mental health scale at age 16 and others have established measurement invariance for the Malaise Inventory at age 42 across these two cohorts,^31^ implying that potential sources of bias such as age effects, survey design, period effects, or cohort specific effects does not influence the way participants in the two cohorts respond to the symptoms described. Moreover, our results indicate that consequences of poor mental health in adolescents are consistently observed between cohorts, and in some cases are more severe in the youngest cohort studied here (e.g. lower likelihood of living with a partner and poorer mental health in midlife). The potential widening of mental health based inequalities in life outcomes, further supports the need to recognise that secular increases in adolescent mental health symptoms is a public health challenge with measurable negative consequences through the lifecourse.^9,13^

In summary, as evidence from more recent generations indicate that common mental health difficulties are increasing further,^9^ our findings indicate that negative consequences of these difficulties might persist or even increase in younger cohorts, with implications not only for social and economic outcomes but also potentially for morbidity and stalling life expectancies.^32^

## Data Availability

The datasets used in this research are available for use to all researchers, free of charge, from the UK data service.

https://ukdataservice.ac.uk/

## Contributors statement

PP conceptualised the study with input from all authors. EJT led on data preparation and analysis. EJT and PP drafted the manuscript and all authors contributed to drafting. All authors approved the final version for submission.

## Acknowledgements

We are grateful for the co-operation and participation of the individuals who voluntarily participate in the 1958 and 1970 birth cohort studies. We thank the Economic and Social Research Council for funding these cohorts through the Centre for Longitudinal Studies (CLS) at the UCL Institute of Education, London. We further thank the Economic and Social Research Council for funding the Cross Cohort Research Programme (CCRP) (grant number: ES/M008584/1).We also thank the Medical Research Council (MRC) (grant numbers: MC_UU_12019/1 and MC_UU_12019//3) for funding Marcus Richards. Dr. Praveetha Patalay is supported by a Wellcome Trust Institutional Strategic Support Fund (ISSF3/ H17RCO/NG1). We would also like to thank a large number of stakeholders from academic, policy-maker and funder communities and colleagues at CLS involved in data collection and management. The funders of the study had no role in study design, data collection, data analysis, data interpretation, or writing of this report. We also thank Prof Alissa Goodman (CLS) for her advice on the economic outcome measures used in this study.

## Data Sharing Statement

The datasets used in this research are available for use to all researchers, free of charge, from the UK data service (https://ukdataservice.ac.uk/).

